# Change in methicillin-resistant *Staphylococcus aureus* testing in the Intensive Care Unit as an antimicrobial stewardship initiative

**DOI:** 10.1101/2022.04.12.22273807

**Authors:** Hayden L. Smith, Samuel P. DuMontier, Amanda M. Bushman, Jonathan R. Hurdelbrink, William J. Yost, Steven R. Craig

**Author notes:** **CORRESPONDING AUTHOR:** Hayden L. Smith, 1415 Woodland Avenue, Suite 140, Des Moines, Iowa, United States;.

## Abstract

**Background:** Methicillin-resistant *Staphylococcus aureus* (MRSA) associated infections are a cause of morbidity/mortality in the Intensive Care Unit (ICU). Vancomycin is an option for treatment but is not without its own risks.

**Purpose:** To institute a testing change to decrease time between ordering of MRSA tests and availability of results in patients admitted to the adult ICU.

**Procedures:** A MRSA testing change was implemented at two adult (i.e., tertiary and community) ICUs located in a U.S. Midwestern health system. The change was implemented in 2018 and included the switch from culture to polymerase chain reaction (PCR) in ICU admitted patients. Study data were collected from 2016-2020 and a Bayesian quantile regression model was fit to examine median level change in time to results and to calculate a counterfactual estimate.

**Main Findings:** During the 58-month period, 71% of 19,975 patients seen at the two ICUs received MRSA testing. In the pre-change period, 91% and 99% of patients at the tertiary and community hospitals received testing via culture, respectively. Culture was used 1% and ∼0% of the time at the hospitals in the post-change period. The counterfactual estimated 36 (95% CrI: 35, 37) and 32 fewer hours (95% CrI: 31, 33) until results were available at the tertiary and community hospital, respectively.

**Conclusions:** Study revealed MRSA results were available in less time at both facilities after testing change. This information can aid anti-microbial stewardship via possibly delaying initiation and/or quicker de-escalation of therapy when results are known.

## INTRODUCTION

Methicillin-resistant *Staphylococcus* aureus infection (MRSA) is a leading cause of severe morbidity, mortality, and economic burden on patients and healthcare systems [Klein]. MRSA colonized patients are commonly present in the Intensive Care Unit (ICU). Between 1992 to 2004, greater than 60% of ICU *S. aureus* infections were attributed to MRSA in the United States (U.S.) and Canada [NNIS]. Between 1995 and 2005, MRSA related hospitalizations nearly doubled, resulting in increasing awareness and need for preventative measures and effective treatment [Klein]. During 2011, the Centers for Disease Control and Prevention estimated 80,461 invasive MRSA infections occurred within the U.S. [Dantes]

Methicillin resistance among *S. aureus* isolates was first identified in the early 1960’s with concomitant, infectious complications arising throughout the decade into the present day [Barber]. MRSA is a common pathogen that is not only part of our own microbiota but also can cause skin/soft tissue, blood stream, bone, heart, and respiratory infections [Lowry, Lakhundi]. It is imperative to initiate empiric antibiotics if clinical suspicion is high for MRSA infection to reduce critical illness as recommended by the Infectious Disease Society of America (IDSA).

Vancomycin is often reflexively utilized as a first choice for empiric antibiotic therapy due to its coverage of MRSA; however, vancomycin therapy comes with its own risks that can disrupt patient care. Acute kidney injury (AKI) is one of the most significant adverse effects of vancomycin. Often, patients in the ICU are susceptible to AKI given numerous potential nephrotoxic agents utilized (i.e., vasopressors, intravenous contrast dye exposure, diuretics, certain beta-lactam antibiotics and aminoglycosides) or comorbidities [Carreno, Rybak, Luther]. In addition, duration of vancomycin exposure, higher doses, and specific patient vulnerability (e.g., previous chronic kidney disease, elevated body mass index, severity of illness, or hemodynamic support) can contribute to nephrotoxicity. Nephrotoxicity has its own multitude of consequences including but not limited to prolonged hospital stay, potential need for dialysis, and increased mortality. [Carreno, Lodise].

It is important to tailor antibiotic regimens to not only deter antibiotic resistance but also limit potential patient harm from unnecessary antibiotic exposure. Antimicrobial stewardship towards MRSA pharmacotherapy is a rapidly developing practice. Early detection of MRSA can help guide and treat acute infection. MRSA polymerase chain reaction (PCR) nasopharyngeal swab testing is one form of rapid detection that has shown excellent performance characteristics especially when detecting pneumonia [Parente DM]. Not only is MRSA-PCR nares screening an effective method for rapid detection but also a potential means for reduction in vancomycin use. [Mergenhagen, Baby N, Woolever] The study objective was to decrease the time between the ordering of a MRSA test and the time when test results are made available in patients being admitted to the adult ICU.

## METHODS

### Study Design and Variables

A multi-site intervention was implemented to change the testing method for MRSA in patients admitted to two adult ICUs within the same health system. The sites included a tertiary hospital (level 1 trauma center; 37 ICU beds) and a community hospital (level IV trauma center; 15 ICU beds) located within 3 miles (Manhattan distance) of each other in the same United States Midwestern city. On February 1, 2018, the laboratories for these facilities changed the MRSA testing default from culture to PCR for patients being admitted to the ICU. Of note, the tertiary hospital used an in-house laboratory and the community hospital used an off-site laboratory. In particular, the community hospital used the tertiary hospital’s laboratory for the first 3.5 years of the study period and then switched to a new off-site laboratory within the health system for the remainder of the reviewed period. Study data was collected for January 1^st^, 2016 through October 31^st^, 2020. This time range included 25 months of pre-change data and 33 months of post-change data.

Test sample collection included a nasopharyngeal swab with cultural test (Becton, Dickinson and Company, Franklin Lakes, New Jersey) based on agar medium to grow *Staph* or PCR test using swab solution based on GeneXpert system (Cepheid, Sunnyvale, California). Test results were added into the labs tab in the electronic health record with no automated notification sent to the ordering provider when results were uploaded into the record. Both tests were available during the two periods, but as noted PCR was deemed the default/preferred method in the post-change period.

Collected data included a hospital indicator (tertiary or community), date and time of admission, date and time of ordered test, test type (culture or PCR), test result (positive or negative), test result date and time, and whether the patient was prescribed vancomycin prior to the posting of the MRSA test result in the medical record (yes or no). An intervention period indicator was constructed (pre-change or post-change period). Collected patient demographic information included age (years) and gender (male or female).

### Data Analysis

Continuous data are reported as medians with interquartile ranges (IQR) and categorical data as counts with percentages. A Bayesian quantile regression model was fit to data to examine the level change in time to lab results at each hospital. Model details are available in the Supplemental File. Results are presented as estimated median times until lab result for pre- and post-change periods and the median difference between these estimates. A counterfactual estimate of change in time to the lab results at the mid-point of the post-change period was constructed (((E(Y^X=0^)), Y: time until results, X: laboratory change [0: represents no laboratory change and 1: represents laboratory change; assuming weak ignorability: X <u>∥</u> Y^x=0^ and X <u>∥</u> Y^x=1^ and a well-defined intervention) and contrasted with the model estimate for that time point (E(Y=*y* | do(X=*x*). This process controlled for changes in time until results were uploaded (i.e., model slopes) within and across study periods. All model-based estimates are reported with 95% credible intervals (CrI) and additional details are available in the Supplemental File. A Monte Carlo (MC) simulation model was fit to quantify the percentage of patients in the post-change period at the tertiary hospital that may probabilistically test positive for MRSA. This estimate represents the hypothetical percentage of positive patients that could have delayed treatment if vancomycin was not initiated until after MRSA results were known. This calculation was based on the empiric number of positive tests in patients receiving vancomycin before MRSA results were known in the post-change period at the tertiary hospital using the beta distribution with 50,000 MC samples. This study received institutional review board approval (IM2017-100).

## RESULTS

### Study sample

Within the 58-month study period, 71% of the 19,975 adult ICU patients received MRSA testing. These patients were distributed with 4,347 and 1,607 in the pre-intervention period and 6,156 and 2,042 in the post-intervention period at the tertiary and community hospitals, respectively. Patients at the tertiary center were 78% male with a median age of 64 (IQR: 52, 74) years, while the community hospital patients were 70% male with a median age of 62 (IQR: 48, 75) years.

### MRSA Testing

In the pre-intervention period 91% and 99% of patients at the tertiary and community hospitals received MRSA testing via culture, respectively. Use of culture testing was 1% and 0% at the tertiary and community hospitals in the post-change period. Across the study periods the MRSA positivity rate was 9.9%, represented by 8.9% and 9.8% in the pre-intervention period and 10.2% and 11.1% in the post-intervention period at the tertiary and community hospitals, respectively. The median time between ICU admitted patient MRSA tests at the tertiary hospital was 2.5 hours and at the community hospital was 8.0 hours across both study periods.

### Time to Test Results

Four (0.002%) MRSA tests were considered outliers and removed from the analytic models since the time until receiving these results were greater than 4 days. Estimated median times to results for the pre- and post-intervention periods by hospital are reported in Table 1 and visualized in Figure 1. Model results revealed an estimated median difference in testing time at the start of the post-intervention period compared to the pre-intervention period to be 38.2 fewer hours (95% CrI: 37.9, 38.5) at the tertiary hospital and 28.8 fewer hours (95% CrI: 28.0, 29.6) at the community hospital. The overall interquartile range value (dispersion) for results in pre-change period at the tertiary and community hospitals were 13- and 12-hours, while these ranges were 1- and 7-hours in the post-change period, respectively. The counterfactual median estimate of difference in time to receiving results at the mid-point in the post-change period was 36.0 fewer hours (95% CrI: 35.4, 36.6) and 31.6 fewer hours (95% CrI: 30.5, 32.8) at the tertiary and community hospitals, respectively. Posterior density plots for model estimates are available in the Supplemental file. Lastly, patients in the pre- versus post-change periods at the tertiary and community hospitals with a vancomycin dose initiated prior to receiving a negative MRSA test were 18% (n=765) versus 17% (n=1,062) and 11% (n=175) versus 14% (n=290), respectively. Based on patient data from the tertiary hospital in the post-change period, if vancomycin was not initiated until the results were known, approximately 16% (95% CI 14%, 18%) of these patients may end up testing positive and having delayed initiation of vancomycin given the drug.

**Table 1.** Median time to receiving methicillin-resistant *Staphylococcus aureus* (MRSA) test results in patients being admitted to an Adult Intensive Care Unit, n=14,148*.

| **Hospital** | **Median Time to MRSA Results in Hours** |  |
| --- | --- | --- |
|  | **Pre-Change** | **Post-Change** |
| Tertiary (Interquartile range) | 41.2 (95% CrI: 41.0, 41.5) | 3.0 (95% CrI: 2.8, 3.1) |
| Community (Interquartile range) | 41.7 (95% CrI: 41.2, 42.2) | 12.9 (95% CrI: 12.2, 13.5) |

CrI: credible interval
\*4 (0.002%) patients dropped from analyses since MRSA test results took longer than 4 days to receive.

**Figure 1:**
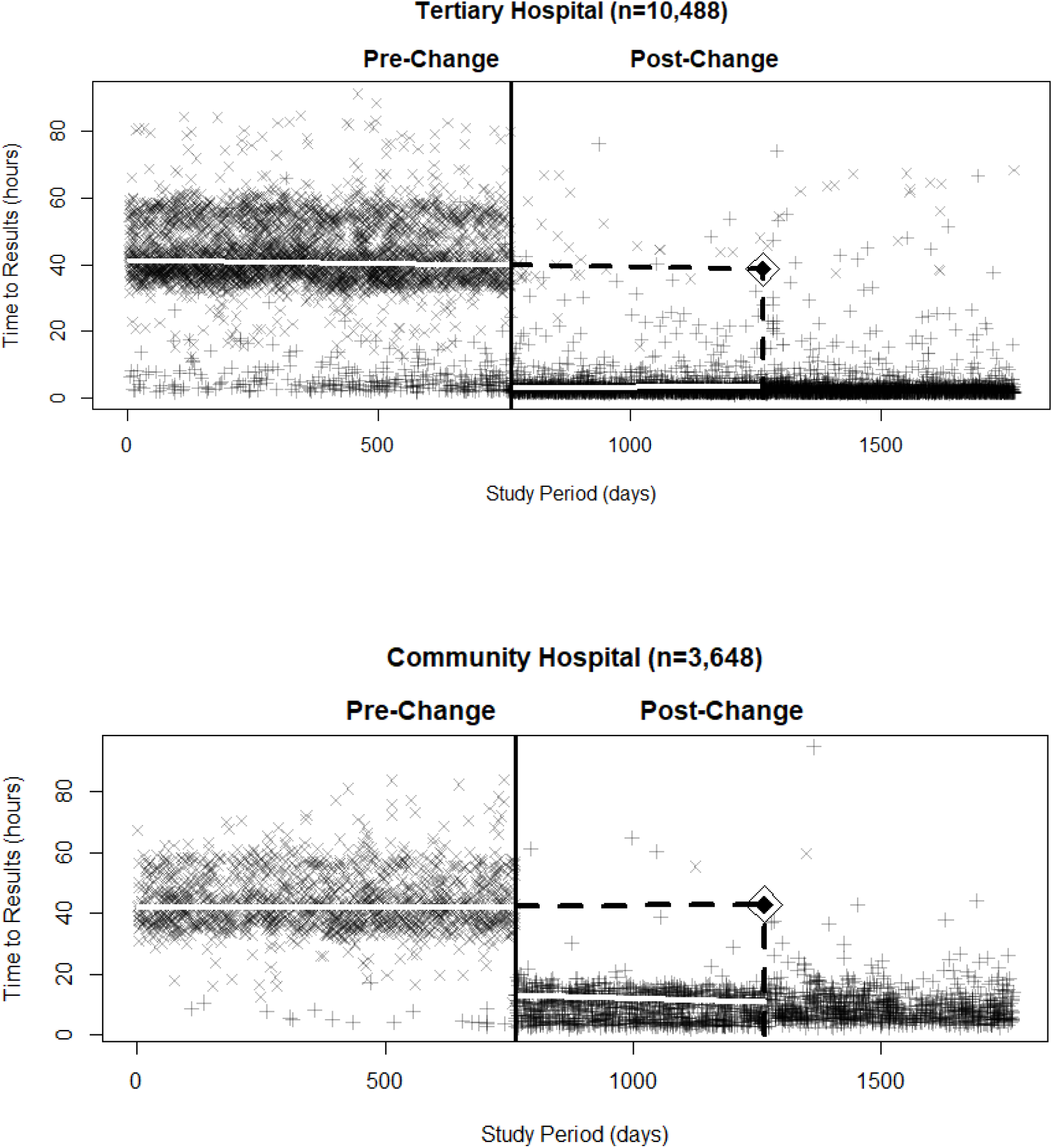
Plotted time (hours) until receiving methicillin-resistant *Staphylococcus aureus* (MRSA) test results in patients admitted to the adult Intensive Care Unit by study period day. Top panel presents tertiary hospital and bottom panel presents community hospital data. Solid black vertical line represents where the test change occurred; two solid white lines are the estimated time to results per period; dashed black horizontal and vertical lines represents the counterfactual estimate at the midpoint of the post- change period with diamond located at the estimate. Symbols: “x” = culture tests; “+” = polymerase chain reaction tests.

## DISCUSSION

The focus of this study was to evaluate time to receiving MRSA results in admitted adult ICU patients after instituting a testing change. At the tertiary hospital the median time to results decreased by over 30 hours. In the community hospital, which used an off-site laboratory, the time decreased by almost 30 hours. Secondary to this change, the number of positive tests went up in the post-change period; however, it is outside the scope of this project to speculate whether this change was related to the testing modality or endemic changes in the population. The variability in time to results decreased in the post-change period at both hospitals compared to the time ranges in the pre-change period.

The presented study reported the number of patients with vancomycin initiated prior to receiving MRSA test results. This group represented patients that could benefit from not having any delays in MRSA testing information. Given a patient’s presentation and pretest probability for MRSA infection, the use of empiric vancomycin may be necessitated in some patients, but potentially delayed until test results are known in other patients given results are known more quickly given PCR. The study revealed results were made available sooner in both hospitals, which can also impact an earlier de-escalation/ discontinuation of antibiotic use in patients testing negative.

Vancomycin therapy has potential detrimental adverse effects that can be both harmful to patients and increase health care costs. Multiple studies have indicated that a MRSA PCR nasopharyngeal swab has a relatively high negative predictive value and is an effective approach to deescalating or discontinuing MRSA antibiotic therapy most notably in patients treated empirically for MRSA pneumonia [Baby N., Woolever, Parente DM]. Moreover, it has been shown that early de-escalation of MRSA antimicrobial therapy did not lead to worse outcomes in such a study sample [Baby N.]. It can be inferred that in situations where MRSA prevalence or overall suspicion for infection is low, avoiding empiric anti-MRSA therapy is plausible until MRSA nasal screening results are verified.

In addition, the post-change study period could have been impacted by COVID-19. The respiratory pandemic led to an increase in mechanical ventilation use which places patients at increased risk for a superimposed infection like MRSA pneumonia. The increase in respiratory illness could have affected data in ways that we are unable to quantify. Additionally, not all ICU patients in the post-change period received a MRSA PCR screen per protocol. Based on study results it can be reasonably inferred that PCR nasal pharyngeal swab is a valuable screening tool to identify MRSA respiratory infection and decrease vancomycin use. It is suspected that the intervention changes made at the study institutions can have lasting implications for each hospital going forward.

### Limitations

In this study the vancomycin courses for patients were not reviewed. The reason for this omission was because initially prescribed vancomycin dosages can vary between patients based on provider preferences and patient presentation. For example, different dosages may have varying coverage and can be titrated to patient based on various factors (e.g., kidney function). Additionally, MRSA test result notifications were not automatically sent to the ordering providers. This communication gap likely slows the process to discontinuation of antibiotics and may vary between providers. This means that MRSA results can be posted in the medical record, but if the provider was not notified of the results, the patient may have remained on the initially prescribed course until it was completed, or the results were realized and acted upon by the provider. Given these issues, it was felt these data may not be completely generalizable to other institutions.

Within the present study there were some patients in the pre-change period that received MRSA testing via PCR while some patients in the post-change period received culture-based testing. The possibility of non-compliance to the preferred testing method per study period could underestimate the optimal theoretical improvement in time to receiving test results that can be realized. This was due to analyses based on an intent-to-treat style design. Though, with the use of the 50^th^ percentile in the quantile regression, the results should not have been overly influenced by a smaller or larger lag in accessibility of results to providers for a minority of patients (i.e., patients getting PCR in the pre-change period or patients getting culture in the post-change period). In the Supplemental file, a deep artificial neural network model was fit to data to show readers what results from an adaptive non-linear model could look like given the increased non-compliance in use of the default testing method.

Patients were not randomized to the testing method given the retrospective sequence design of the study. This could result in making the exchangeability of patient characteristics between periods possibly unbalanced. Though, theoretically a study of a testing and laboratory change would occur in a standardized fashion and negate any most concerns about patient differences associated with the testing method used. Lastly, the study did not have a staggered roll-out across the hospitals or include a negative control group to help understand the potential for an unknown historical bias occurring at the time of the testing change. This concern was indirectly addressed by providing an extended pre- and post-change time series, which revealed no apparent occurrences of exogenous shocks or slope changes in the series beyond the study intervention point.

## CONCLUSIONS

The study revealed a decrease in time to MRSA test results and a decrease in the variability of these times in the post-change period. The study design replicated the intervention at two different types of facilities in order to show the potential generalizability of the change and to corroborate findings.

## Supporting information

SQUIRE Checklist

## Data Availability

All data produced in the present study are not available upon request to the authors.

## ACKNOWLEDGEMENTS

David J. Aman, Chanteau M. Ayers, Austin J. Boeckman, Frank J. Caligiuri, Brook N. Delpierre, Laura C. Elliott, Julie A. Gibbons, Vali P. Potter, Rossana M. Rosa.

