## Supplemental File for "Change in methicillin-resistant *Staphylococcus aureus* testing in the Intensive Care Unit as an antimicrobial stewardship initiative"

***Statistical Model***

The a priori analytic plan was to examine data and fit a Bayesian interrupted time series and report estimated medians and a counterfactual. The study model was conducted using runjags (2.2.0-2) in R version 4.0.2 (2020-06-22). Model chains for estimates were initiated based on the empiric median time to result values for the two study periods per hospital using study data and a random draw from the uniform distribution for sigma and rho (i.e., sigma = runif(1, 0.1, 10); rho = runif(1, -1, 1)). Priors used in the models are presented below.

Tertiary hospital:

Pre-change intercept ~ dnorm(40, 2)

Pre-change slope ~ dnorm(0, 1)

Post-change intercept ~ dgamma(5, 2)

Post-change slope ~ dnorm(0, 1)

Sigma ~ dunif(0.1, 10)

Rho ~ dunif(-1, 1)

Community hospital:

Pre-change intercept ~ dnorm(40, 2)

Pre-change slope ~ dnorm(0, 1)

Post-change intercept ~ dnorm(8, 2)

Post-change slope ~ dnorm(0, 1)

Sigma ~ dunif(0.1, 10)

Rho ~ dunif(-1, 1)

An examination of the above model revealed patients in the pre-change and post-change periods could have received the non-default test (i.e., PCR in lieu of culture and vice versa). These 'non-compliant' tests represented a more complex underlying data generating process where the two tests were influencing the dependent variable estimates in the model within both periods. This was also seen in a multimodal residual distribution for the model errors. The existence of non-compliance also resulted in predictions of estimates varying based on what the previous test was for that prediction. For example, if the prior test was culture with a 44-hour result time and the next test was PCR, predicted estimates would be a weighted value around 16-hours until results and not near the actual value of around 4-hours at the tertiary hospital. Also, if the next test was a culture its predicted estimate was also a weighted version based now on the PCR test. This resulted in a varying identification of the estimated outcome. Secondarily, autocorrelation function and partial autocorrelation function plots revealed a lack of correlation patterns in model errors across time.

Given the purpose of the study was to model the median change in time to results between the study periods, a Bayesian quantile regression model was selected to examine the primary objective of the study, since it would be able to provide a robust estimate of the median. The bayesQR (2.3) R package was used to fit study data in a quantile regression using an intercept, pre-change period slope, post-change period indicator, and post-change slope (interaction term) variable with 20,000 samples with a 100 burn-in, thinning=2, and chains=2. Priors for the model are presented below. Of note, bayesQR uses asymmetric Laplace densities which restricted prior distribution options, which meant gamma priors could no longer be used).

Tertiary hospital:

Pre-change intercept ~ dnorm(40, 2)

Pre-change slope ~ dnorm(0, 1)

Post-change intercept ~ dnorm(3, 0.4)

Post-change slope ~ dnorm(0, 1)

Sigma ~ default flat prior

Community hospital:

Pre-change intercept ~ dnorm(40, 2)

Pre-change slope ~ dnorm(0, 1)

Post-change intercept ~ dnorm(9, 2)

Post-change slope ~ dnorm(0, 1)

Sigma ~ default flat prior

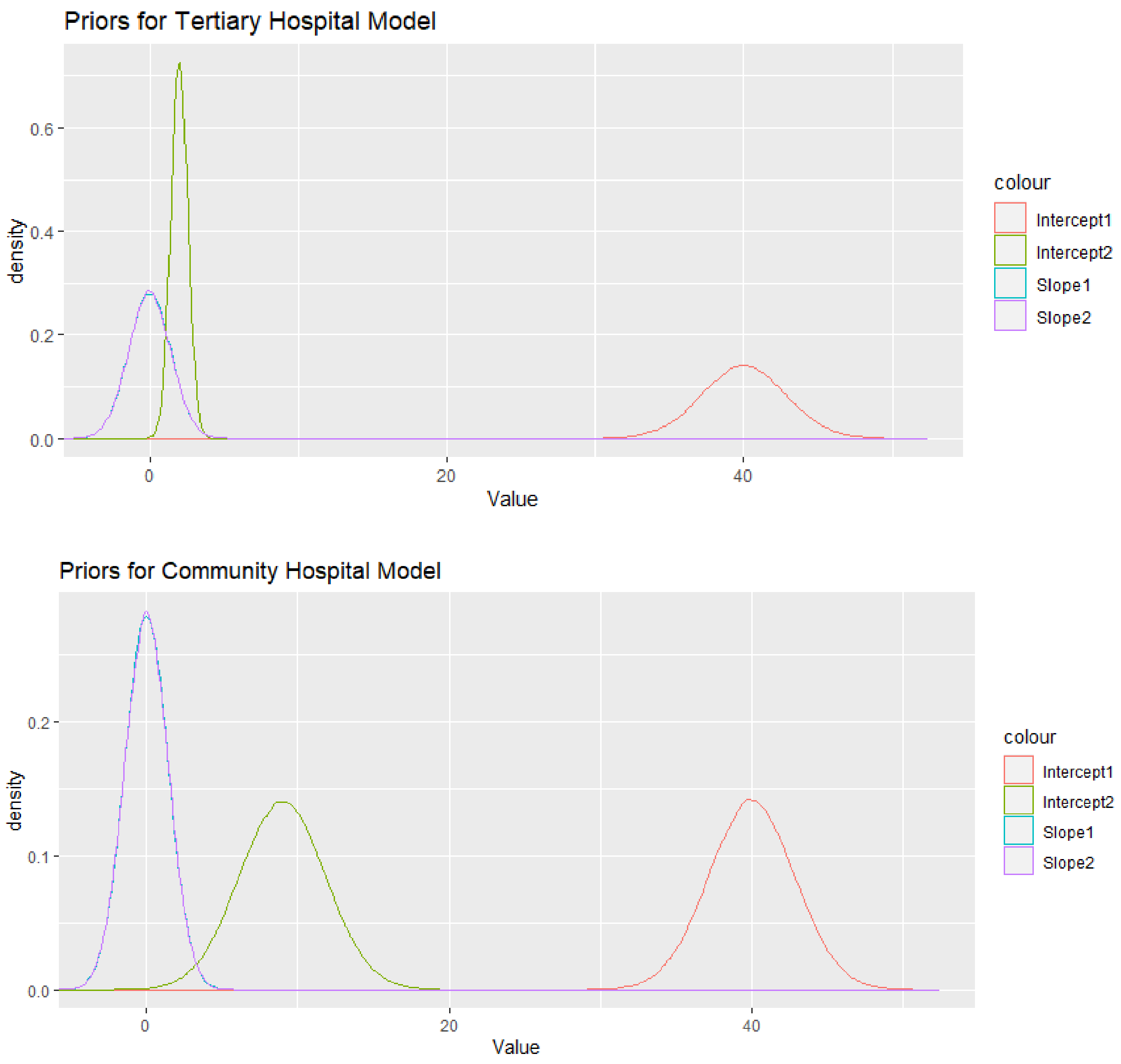


**Figure 1.** Top panel presents the priors used in the tertiary hospital model and the bottom panel presents the priors used in the community hospital model. Priors for slope values for the two-line segments were the same and mostly overlap in the above figures.

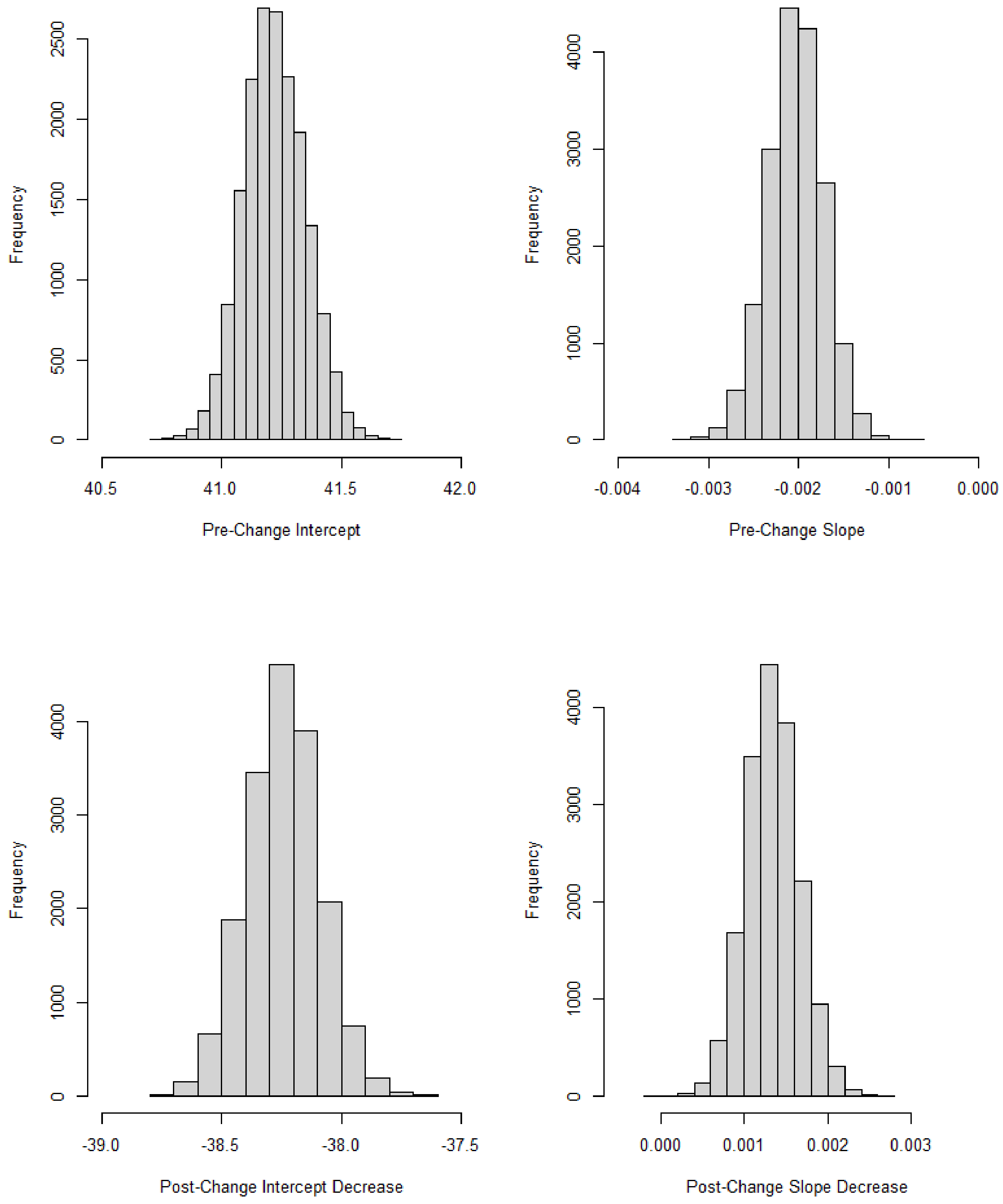


**Figure 2.** Tertiary hospital posterior estimates; x-axes represent hours related to receiving methicillin-resistant *Staphylococcus aureus* (MRSA) results with the pre-change period being before the testing switch from culture to polymerase chain reaction in the adult Intensive Care Unit.

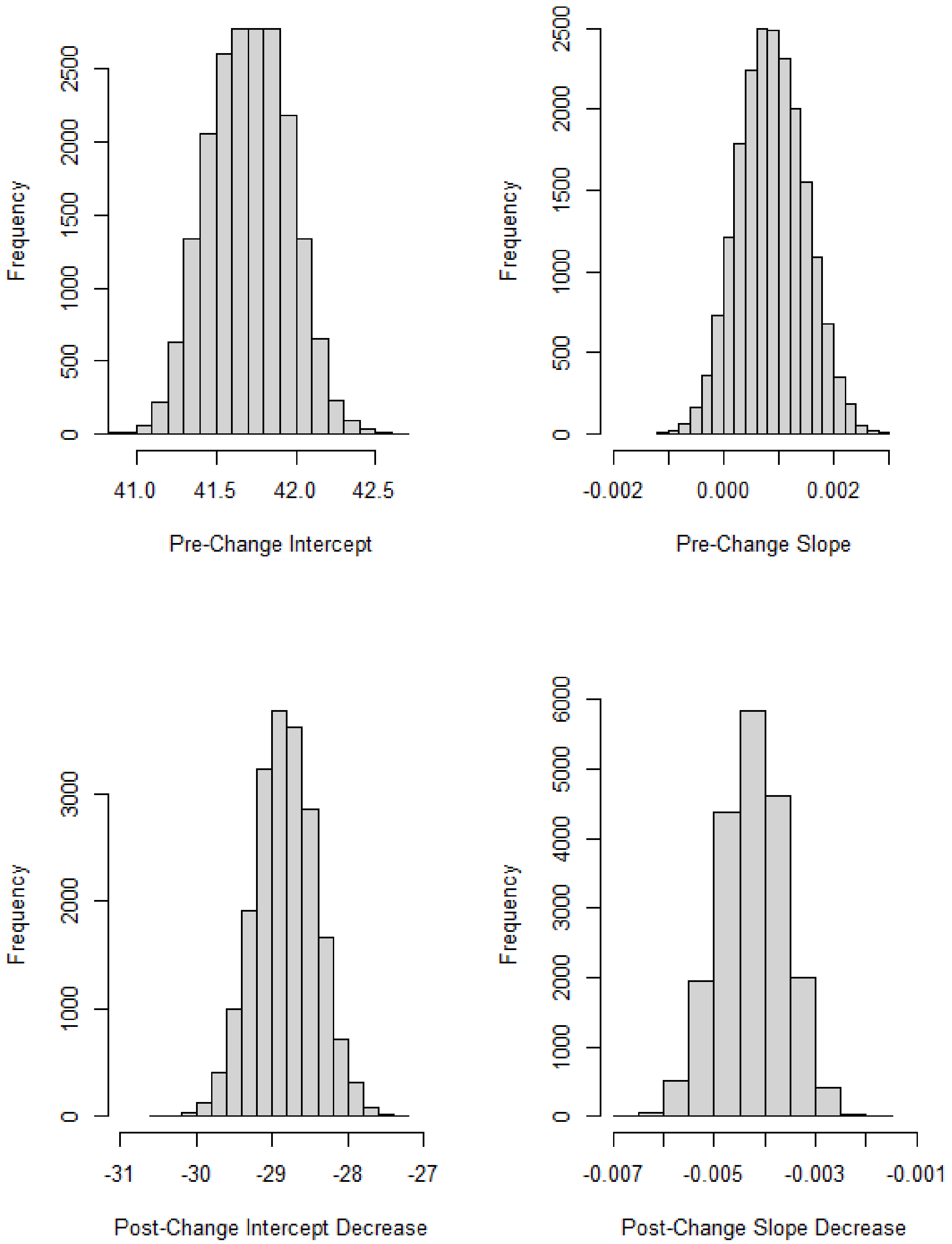


**Figure 3.** Community hospital posterior estimates; x-axes represent hours related to receiving methicillin-resistant *Staphylococcus aureus* (MRSA) results with the pre-change period being before the testing switch from culture to polymerase chain reaction in the adult Intensive Care Unit.

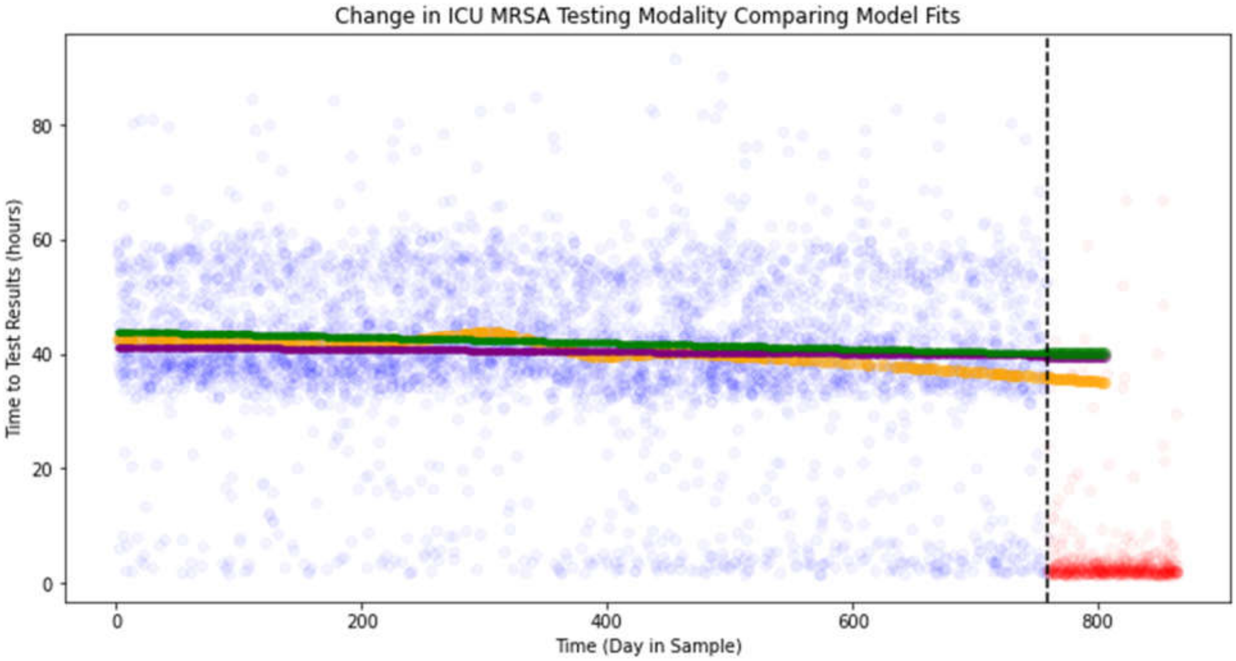


**Figure 4.** Presented are two alternative model fits. The green line represents the quantile regression from the paper. The purple line is a robust regression (i.e., Huber), while the orange line is a deep artificial neural network (i.e., three hidden layers with reLu activations, initial null bias and Glorot normal weights, batch normalization, and 10% dropout). The neural network shows what a more adaptive non-linear prediction can look like given the presence of increased non-compliance in default testing method near the end of the study period (i.e., dashed vertical line).
