## Supplementary material for "Change in methicillin-resistant *Staphylococcus aureus* testing in the Intensive Care Unit as an antimicrobial stewardship initiative": SQUIRE Checklist

### SQUIRE (Standards for QUality Improvement Reporting Excellence) Checklist

Final revision – 4-29-08

- These guidelines provide a framework for reporting formal, planned studies designed to assess the nature and effectiveness of interventions to improve the quality and safety of care.
- It may not be possible to include information about every numbered guideline item in reports of original formal studies, but authors should at least consider every item in writing their reports.
- Although each major section (i.e., Introduction, Methods, Results, and Discussion) of a published original study generally contains some information about the numbered items within that section, information about items from one section (for example, the Introduction) is often also needed in other sections (for example, the Discussion).

| Text Section; Item Number and Name | Section or Item Description | Reported (Yes/No) | Reported on Page No. |
|---|---|---|---|
| **Title and Abstract** | *Did you provide clear and accurate information for finding, indexing, and scanning your paper?* | | |
| 1. Title | a. Indicates the article concerns the improvement of quality (broadly defined to include the safety, effectiveness, patient-centeredness, timeliness, efficiency, and equity of care) | Yes | |
| | b. States the specific aim of the intervention | Yes | |
| | c. Specifies the study method used (for example, "A qualitative study," or "A randomized cluster trial") | Yes | |
| 2. Abstract | Summarizes precisely all key information from various sections of the text using the abstract format of the intended publication | Yes | |
| **Introduction** | *Why did you start?* | | |
| 3. Background Knowledge | Provides a brief, non-selective summary of current knowledge of the care problem being addressed, and characteristics of organizations in which it occurs | Yes | |
| 4. Local problem | Describes the nature and severity of the specific local problem or system dysfunction that was addressed | Yes | |
| 5. Intended improvement | a. Describes the specific aim (changes/improvements in care processes and patient outcomes) of the proposed intervention | Yes | |
| | b. Specifies who (champions, supporters) and what (events, observations) triggered the decision to make changes, and why now (timing) | Yes | |
| 6. Study question | States precisely the primary improvement-related question and any secondary questions that the study of the intervention was designed to answer | Yes | |
| **Methods** | *What did you do?* | | |
| 7. Ethical issues | Describes ethical aspects of implementing and studying the improvement, such as privacy concerns, protection of participants' physical well-being, and potential author conflicts of interest, and how ethical concerns were addressed | Yes | |
| 8. Setting | Specifies how elements of the environment considered most likely to influence change/improvement in the involved site or sites were identified and characterized | Yes | |

| Text Section; Item Number and Name | Section or Item Description | Reported (Yes/No) | Reported on Page No. |
|---|---|---|---|
| 9. Planning the intervention | a. Describes the intervention and its component parts in sufficient detail that others could reproduce it | Yes | |
| | b. Indicates main factors that contributed to choice of the specific intervention (for example, analysis of causes of dysfunction; matching relevant improvement experience of others with the local situation) | Yes | |
| | c. Outlines initial plans for how the intervention was to be implemented: e.g., what was to be done (initial steps; functions to be accomplished by those steps; how tests of change would be used to modify intervention), and by whom (intended roles, qualifications, and training of staff) | Yes | |
| 10. Planning the study of the intervention | a. Outlines plans for assessing how well the intervention was implemented (dose or intensity of exposure) | Yes | |
| | b. Describes mechanisms by which intervention components were expected to cause changes, and plans for testing whether those mechanisms were effective | Yes | |
| | c. Identifies the study design (for example, observational, quasi-experimental, experimental) chosen for measuring impact of the intervention on primary and secondary outcomes, if applicable | Yes | |
| | d. Explains plans for implementing essential aspects of the chosen study design, as described in publication guidelines for specific designs, if applicable (see, for example, http://www.equator-network.org) | Yes | |
| | e. Describes aspects of the study design that specifically concerned internal validity (integrity of the data) and external validity (generalizability) | Yes | |
| 11. Methods of evaluation | a. Describes instruments and procedures (qualitative, quantitative, or mixed) used to assess a) the effectiveness of implementation, b) the contributions of intervention components and context factors to effectiveness of the intervention, and c) primary and secondary outcomes | Yes | |
| | b. Reports efforts to validate and test reliability of (11.b. *cont.*) assessment instruments | Yes | |
| | c. Explains methods used to assure data quality and adequacy (for example, blinding; repeating measurements and data extraction; training in data collection; collection of sufficient baseline measurements) | Yes | |
| 12. Analysis | a. Provides details of qualitative and quantitative (statistical) methods used to draw inferences from the data | Yes | |
| | b. Aligns unit of analysis with level at which the intervention was implemented, if applicable | Yes | |
| | c. Specifies degree of variability expected in implementation, change expected in primary outcome (effect size), and ability of study design (including size) to detect such effects | Yes | |

| Text Section; Item Number and Name | Section or Item Description | Reported (Yes/No) | Reported on Page No. |
|---|---|---|---|
| | d. Describes analytic methods used to demonstrate effects of time as a variable (for example, statistical process control) | Yes | |
| **Results** | *What did you find?* | | |
| 13. Outcomes | | Yes | |
| 13a. Nature of setting and improvement intervention | i) Characterizes relevant elements of setting or settings (for example, geography, physical resources, organizational culture, history of change efforts), and structures and patterns of care (for example, staffing, leadership) that provided context for the intervention | Yes | |
| | ii) Explains the actual course of the intervention (for example, sequence of steps, events or phases; type and number of participants at key points), preferably using a time-line diagram or flow chart | Yes | |
| | iii) Documents degree of success in implementing intervention components | Yes | |
| | iv) Describes how and why the initial plan evolved, and the most important lessons learned from that evolution, particularly the effects of internal feedback from tests of change (reflexiveness) | Yes | |
| 13b. Changes in processes of care and patient outcomes associated with the intervention | i) Presents data on changes observed in the care delivery process | Yes | |
| | ii) Presents data on changes observed in measures of patient outcome (for example, morbidity, mortality, function, patient/staff satisfaction, service utilization, cost, care disparities) | Yes | |
| | iii) Considers benefits, harms, unexpected results, problems, failures | Yes | |
| | iv) Presents evidence regarding the strength of association between observed changes/improvements and intervention components/context factors | Yes | |
| | v) Includes summary of missing data for intervention and outcomes | Yes | |
| **Discussion** | *What do the findings mean?* | | |
| 14. Summary | a) Summarizes the most important successes and difficulties in implementing intervention components, and main changes observed in care delivery and clinical outcomes | Yes | |
| | b) Highlights the study's particular strength | Yes | |
| 15. Relation to other evidence | Compares and contrasts study results with relevant findings of others, drawing on broad review of the literature; use of a summary table may be helpful in building on existing evidence | Yes | |
| 16. Limitations | a. Considers possible sources of confounding, bias, or imprecision in design, measurement, and analysis that might have affected study outcomes (internal validity) | Yes | |

| Text Section; Item Number and Name | Section or Item Description | Reported (Yes/No) | Reported on Page No. |
|---|---|---|---|
| | b. Explores factors that could affect generalizability (external validity), for example: representativeness of participants; effectiveness of implementation; dose-response effects; features of local care setting | Yes | |
| | c. Addresses likelihood that observed gains may weaken over time, and describes plans, if any, for monitoring and maintaining improvement; explicitly states if such planning was not done | Yes | |
| | d. Reviews efforts made to minimize and adjust for study limitations | Yes | |
| | e. Assesses the effect of study limitations on interpretation and application of results | Yes | |
| 17. Interpretation | a. Explores possible reasons for differences between observed and expected outcomes | Yes | |
| | b. Draws inferences consistent with the strength of the data about causal mechanisms and size of observed changes, paying particular attention to components of the intervention and context factors that helped determine the intervention's effectiveness (or lack thereof), and types of settings in which this intervention is most likely to be effective | Yes | |
| | c. Suggests steps that might be modified to improve future performance | Yes | |
| | d. Reviews issues of opportunity cost and actual financial cost of the intervention | Yes | |
| 18. Conclusions | a. Considers overall practical usefulness of the intervention | Yes | |
| | b. Suggests implications of this report for further (18.b. *cont.*) studies of improvement interventions | Yes | |
| **Other Information** | *Were other factors relevant to conduct and interpretation of the study?* | | |
| 19. Funding | Describes funding sources, if any, and role of funding organization in design, implementation, interpretation, and publication of study | Yes | |

**Once you have completed this checklist, please save a copy and upload it as part of your submission. DO NOT include this checklist as part of the main manuscript document. It must be uploaded as a separate file.**
